## Supplemental figures for "Malaria surveillance reveals parasite relatedness, signatures of selection, and correlates of transmission across Senegal" (from another version of this preprint: version 1, 2023-04-17)

a

e

c

d

b

*Supplemental Figure 1*. *Steps in selecting SNP sites for analysis, based on the complete Pf3K dataset. a) Mean rate of heterozygous calls for nonoverlapping 2 kb windows across the* P. falciparum *genome. b) Same as (a) but restricted to ‘core’ regions of the genome [1]. c) Distribution of bin heterozygosities with and without non-core regions. d) Heterozygosity as a function of distance from known indels; ‘0’ bin contains sites within indels. e) Rate of heterozygous calls after application of all filters.*

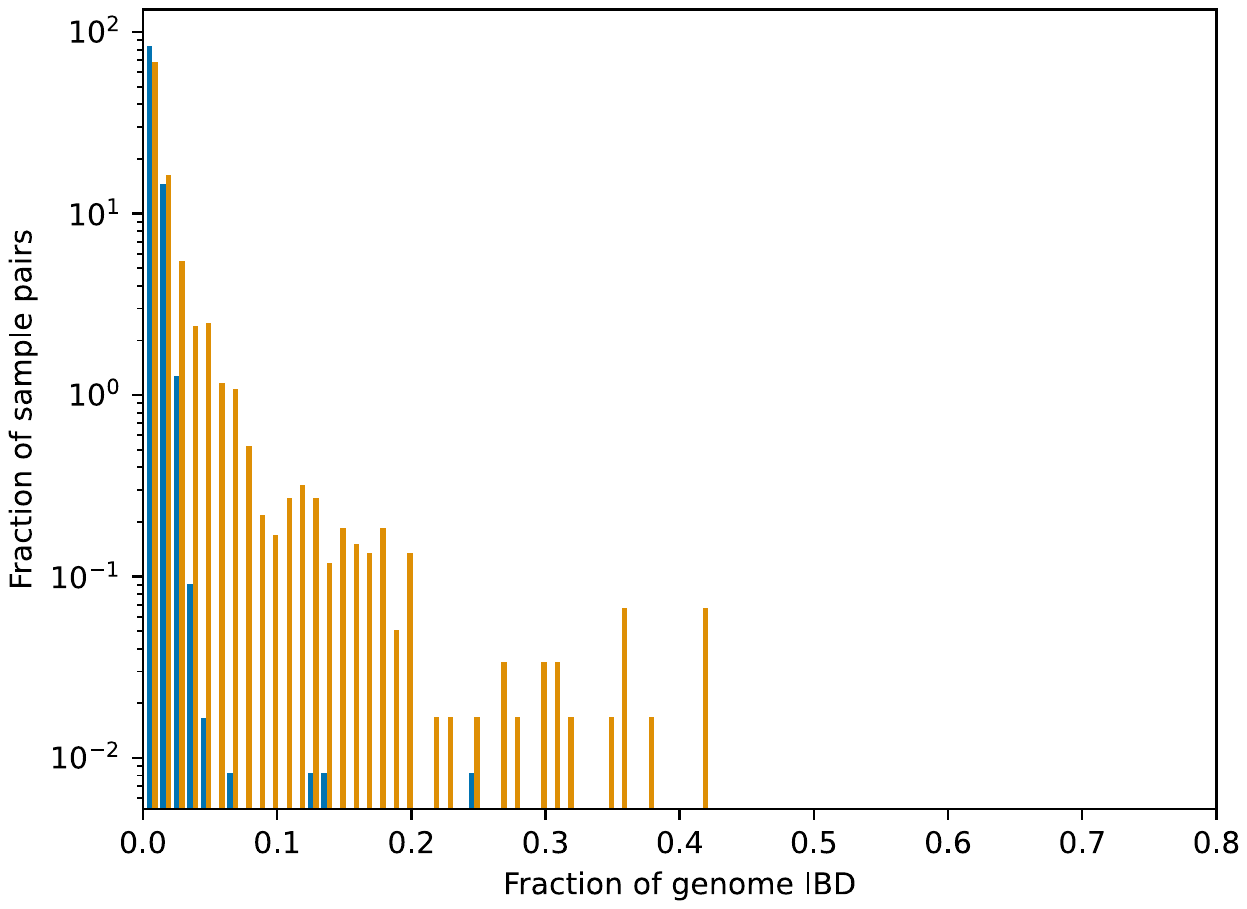

*Supplemental Figure 2.* *IBD fraction for sample pairs enriched (orange) and depleted (blue) for relatedness; only higher coverage samples shown.*

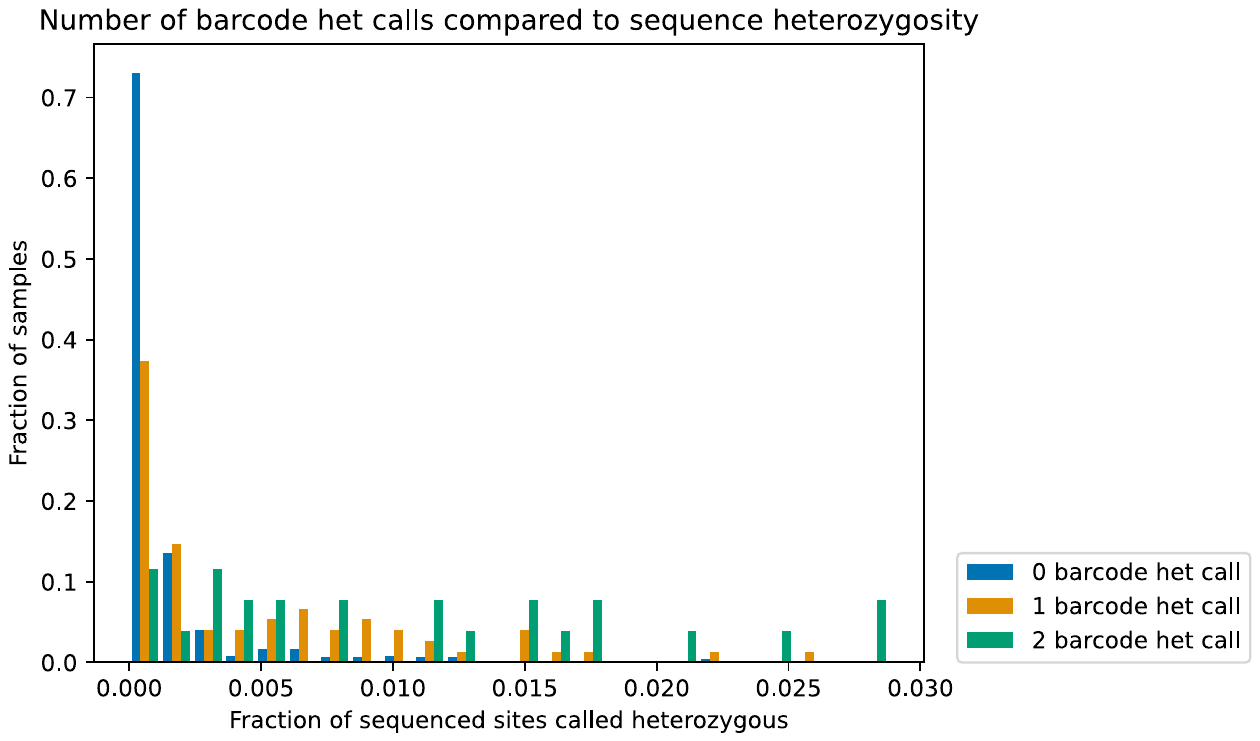

*Supplemental Figure 3. Comparison of complexity of infection measures from barcode and sequence data. Shown is the distribution of sequenced sites that were called as heterozygous for samples with both barcode and whole sequence data. Samples with 0 or 1 heterozygous barcode sites were classified as probable monogenomic.*

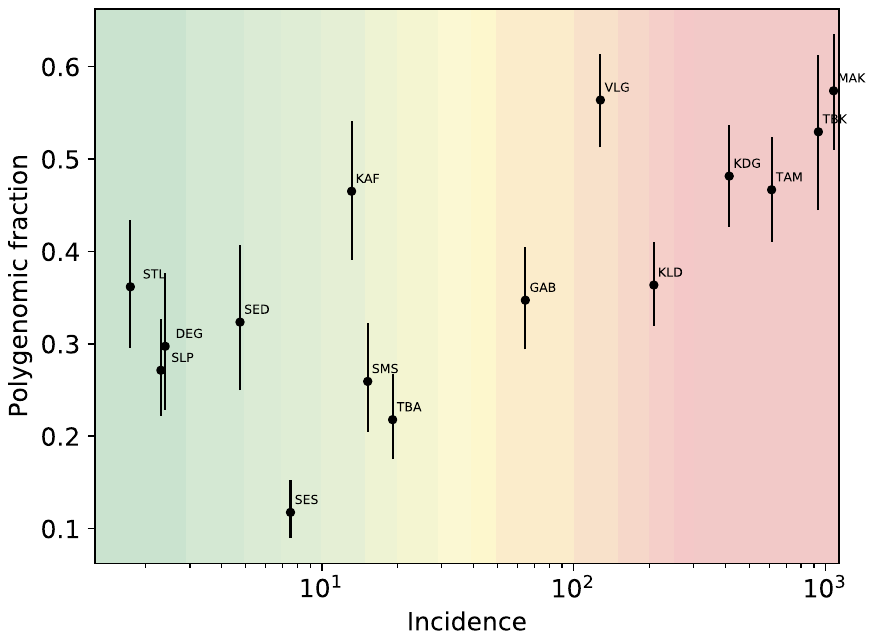

*Supplemental Figure 4. Polygenomic fraction vs reported incidence (per 1000 per year) at study site (log scale).*

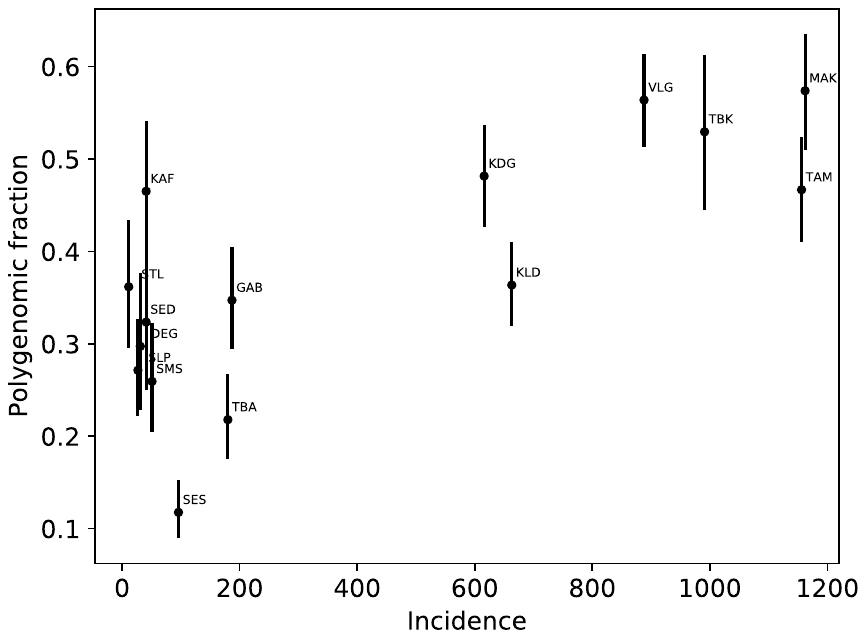

*Supplemental Figure 5. Polygenomic fraction as a function of estimated incidence (per 1000 per year), where the incidence has been adjusted for care seeking, testing of suspected cases, and reporting completeness.*

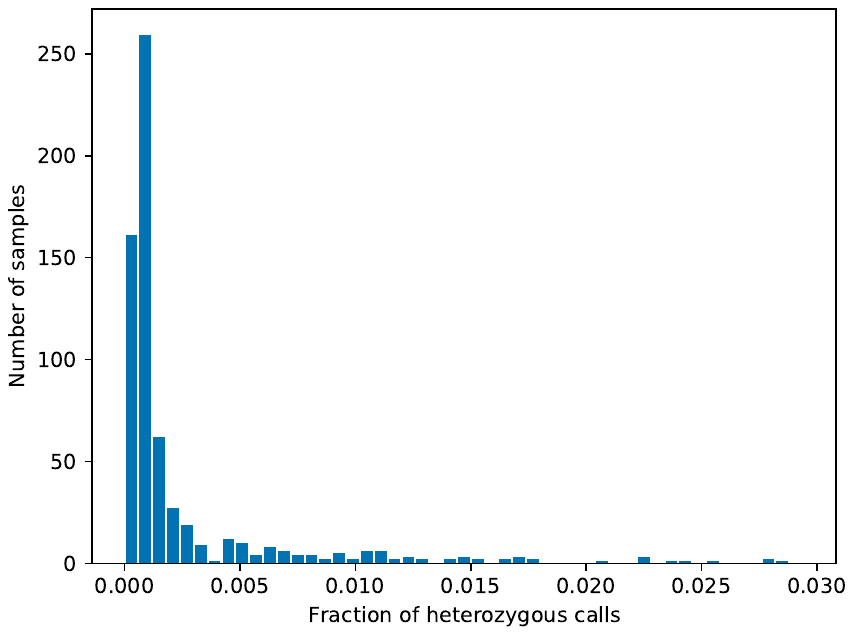

*Supplemental Figure 6. Fraction of heterozygous genotypes (out of all called sites) for all sequenced samples. Samples with a fraction > 0.0024 were considered possibly polygenomic and excluded from further analysis.*

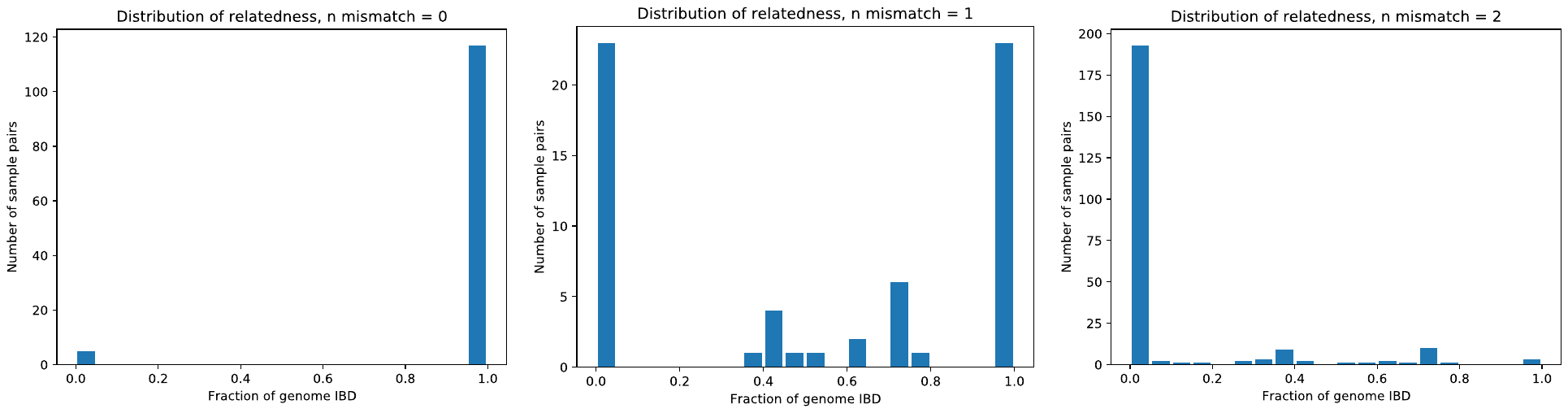

*Supplemental Figure 7. Accuracy of relatedness inference from barcode data. The panels show the distribution of genome wide IBD fraction as determined from sequence data for sample pairs with 0, 1, and 2 barcode mismatches, respectively,*

| Genetic measures in model | R^2^ | Adjusted R^2^ |
| --- | --- | --- |
| polygenomic fraction (poly) | 0.564 | 0.527 |
| total relatedness (total) | 0.194 | 0.127 |
| poly + total | 0.619 | 0.550 |
| poly + clonal | 0.590 | 0.515 |
| poly + partial | 0.593 | 0.519 |
| poly + clonal + partial | 0.621 | 0.507 |

*Supplemental Table 1. Performance of different genetic measures as predictors of malaria incidence at each site, as determined by regressing log(incidence) on the indicated measures. Clonal: clonal relatedness; partial: partial relatedness.*

**Supplemental Information: IBD pileup plots for all chromosomes**

*The fraction of sample pairs that are IBD as a function of genomic position, split into samples that are (“within cluster”) or are not (“outside cluster”) part of the large network of related parasites. Individual chromosomes are displayed (chromosome 1 – 14) with the position of some individual genes identified by green dotted line including: dhfr (Pfdhfr, chromosome 4), mdr1 (Pfmdr1, chromosome 5), aat1 (Pfaat1, chromosome 6), crt (Pfcrt, chromosome 7), dhps (Pfdhps, chromosome 8), gch1 (Pfgch1, chromosome 12), and fd (ferredoxin, chromosome 13).*

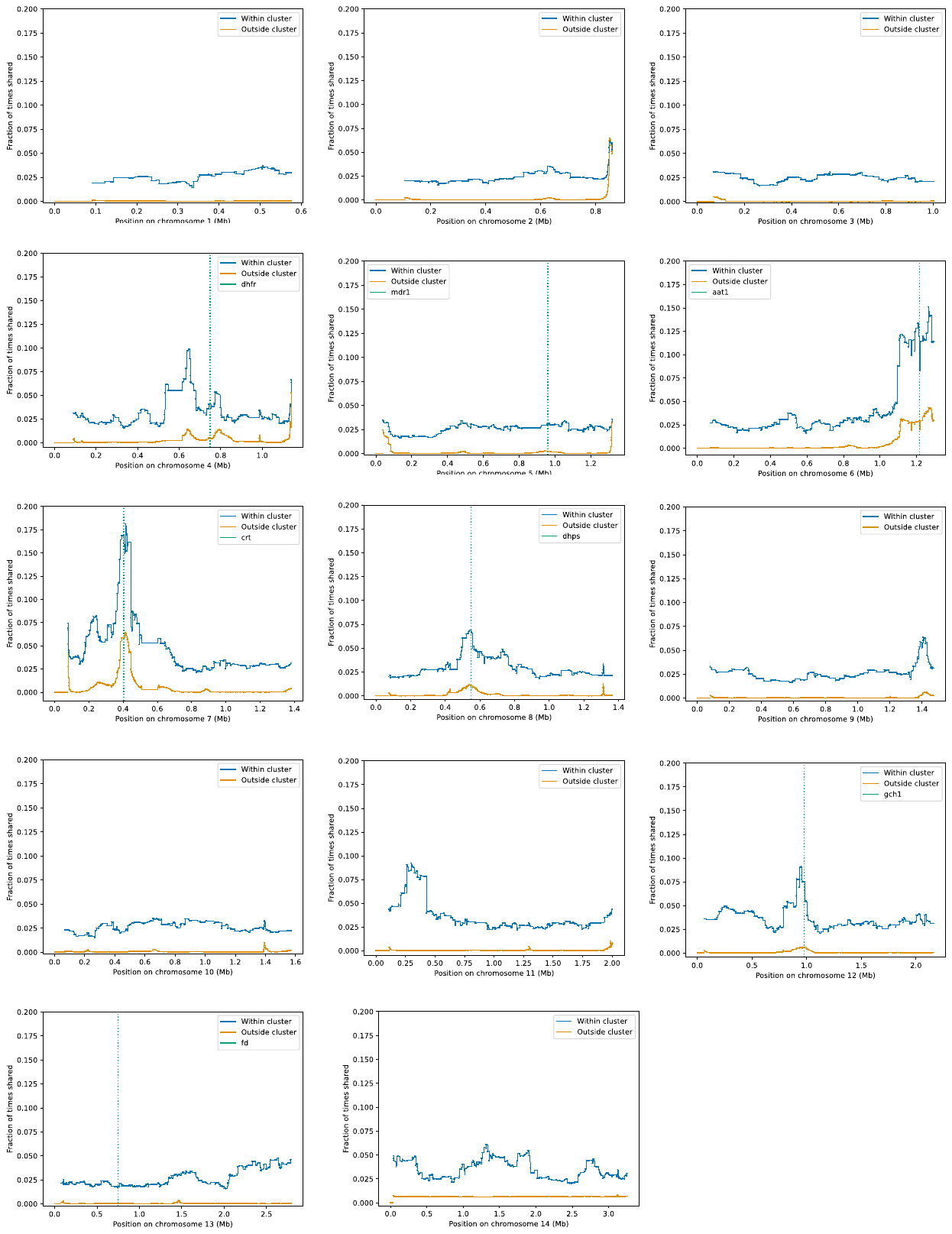
